# Association between COVID-19 infection rates by region and implementation of non-pharmaceutical interventions: A cross-sectional study in Japan

**DOI:** 10.1101/2021.07.26.21261107

**Authors:** Tomomi Anan, Tomohiro Ishimaru, Ayako Hino, Tomohisa Nagata, Seiichiro Tateishi, Mayumi Tsuji, Akira Ogami, Shinya Matsuda, Yoshihisa Fujino, for the CORoNaWork project

**Affiliations:** University of Occupational and Environmental Health, Japan

## Abstract

**Background:** During a pandemic, non-pharmaceutical interventions (NPIs) play an important role in protecting oneself from infection and preventing the spread of infection to others. There are large regional differences in COVID-19 infection rates in Japan. We hypothesized that the local infection incidence may affect adherence to individual NPIs.

**Methods:** This cross-sectional study was conducted online among full-time workers in Japan in December 2020. Data from a total of 27,036 participants were analyzed. The questionnaire asked the respondents to identify their habits regarding seven well-known NPIs.

**Results:** Compared to the region with the lowest infection rate, the odds ratios for the region with the highest infection rate were 1.24 (p<0.001) for wearing a mask in public, 1.08 (p=0.157) for washing hands after using the bathroom, 1.17 (p=0.031) for disinfecting hands with alcohol sanitizers when entering indoors, 1.54 (p<0.001) for gargling when returning home, 1.45 (p<0.001) for ventilating the room, 1.33 (p<0.001) for disinfecting or washing hands after touching frequently touched surfaces, and 1.32 (p<0.001) for carrying alcohol sanitizers when outdoors. Five of the seven NPIs showed statistically significant trends across regional infection levels, the two exceptions being wearing a mask in public and washing hands after using the bathroom. Multivariate adjustment did not change these trends.

**Conclusions:** This study found that NPIs were more prevalent in regions with higher incidence rates of COVID-19 in Japanese workers. The findings suggest that the implementation of NPIs was influenced not only by personal attributes but also by contextual effects of the local infection level.

## Introduction

COVID-19 has spread rapidly all around the world since December 2019. The World Health Organization (WHO) declared COVID-19 a pandemic on March 11, 2020 (1). Various efforts have been made at a policy level in many countries to prevent the spread of infection during the pandemic, which is ongoing. Many countries have implemented lockdowns, curfews of restaurants and bars, physical distancing, bans on social gatherings, and school closures. By the end of 2020, vaccines against COVID-19 had been developed and their roll out had commenced. The Japanese government declared a “state of emergency” in response to record numbers of cases, and asked the public to refrain from going out, close restaurants or restrict their opening times, work remotely, and limit or cancel events (2).

During a pandemic, non-pharmaceutical interventions (NPIs) play an important role in protecting oneself from infection and preventing the spread of infection to others. NPIs are defined as behaviors or actions that individuals or communities can take to slow the spread of pathogens, such as washing hands, wearing masks, and physical distancing; they do not include taking medication or receiving vaccines (3). Centers for disease control and prevention (CDCs) recommend wearing a mask, staying at least six feet away from others, avoiding crowds and poorly ventilated spaces, washing hands frequently, cleaning and disinfecting frequently touched surfaces, and monitoring one’s health daily (4). The Japanese government has called on the public to focus on avoiding the “three Cs”: closed spaces, crowded places, and close-contact settings, and has reminded people to wash their hands, wear masks, ventilate their houses, and avoid eating out with others (5).

Although the implementation of NPIs is important for preventing the spread of infection, some people do not follow the recommended interventions, and some even oppose them. It is already known that adherence to individual NPIs is influenced by demographic factors such as gender, age, and place of residence (6); social factors such as education, income (7,8), and sources of information (9); and psychological factors such as anxiety, fear, political ideology, and health beliefs (7,8,10,11). In the U.S., many people in urban areas resist wearing masks, partly due to political ideology (6,12). Unlike in other countries, the Japanese government’s infection control actions do not include mandatory measures such as lockdowns, but mostly request cooperation from the public. Therefore, individuals’ attitudes toward the NPIs is a key issue for combatting the spread of infection.

There are large regional differences in COVID-19 infection rates in Japan, which may lead to differences in people’s risk perception. The risk perception regarding COVID-19 differs widely among places and individuals, and can affect the spread of the virus (13). One study reported that Japanese people placed most trust in local information (9) about infection status and hospital bed occupancy rates. As the infection rate in a region rises, people start to hear about cases close to them, and infection prevention measures are strengthened in workplaces and public places. We assume that people’s risk perceptions are influenced by contextual and environmental factors.

We hypothesized that the local infection incidence rate may affect adherence to individual NPIs. We considered that the incidence rate of COVID-19 in a given region is related to the implementation of such interventions through people’s risk perception. To our knowledge, however, very few reports have examined the rate of implementation of NPIs in relation to local infection rates. Here, to test this hypothesis, we examined the association between regional differences in COVID-19 infection levels in Japan and individual NPIs.

## Methods

### Study Design

We conducted a survey from December 22 to 26, 2020, during the third wave of the COVID-19 pandemic, as a part of the Collaborative Online Research on the Novel-coronavirus and Work (CORoNaWork) Project (14). This cross-sectional study was conducted online among full-time workers in Japan.

The study was approved by the ethics committee of the University of Occupational and Environmental Health, Japan (reference No. R2-079 and R3-006). Participants provided informed consent by completing a form on the survey website.

### Study Population

A total of 33,087 participants answered an online, self-administered questionnaire. After excluding invalid responses, 27,036 were eligible for analysis. The age-range of the target population was 20 to 65 years. To take account of regional characteristics, the 47 Japanese prefectures were classified into four levels based on the level of infection. Completed questionnaire samples were extracted from these four regions to obtain equal sample sizes.

Region 1 consisted of Fukushima, Yamaguchi, Aomori, Ehime, Yamagata, Nagasaki, Iwate, Tokushima, Shimane, Kagawa, Niigata, Tottori, and Akita prefectures; region 2 of Nagano, Saga, Tochigi, Oita, Toyama, Okayama, and Fukui prefectures; region 3 of Gunma, Ishikawa, Gifu, Kumamoto, Ibaraki, Miyagi, Hiroshima, Shiga, Mie, Kochi, Shizuoka, Wakayama, Miyazaki, Yamanashi, and Kagoshima prefectures; and region 4 of Tokyo, Kanagawa, Saitama, Chiba, Okinawa, Osaka, Hokkaido, Aichi, Hyogo, Fukuoka, Kyoto, and Nara prefectures.

### Assessment of non-pharmaceutical interventions

The questionnaire asked respondents to report on their habits regarding seven types of NPIs in the last month, namely wearing a mask in public, washing hands after using the bathroom, disinfecting hands with alcohol sanitizers when going inside, gargling when returning home, opening windows or doors to ventilate the room, disinfecting or washing hands after touching frequently touched surfaces such as doorknobs or railings, and carrying alcohol sanitizers when going out. Participants answered from the following options: “always do”, “mostly do”, “not often”, or “almost never”.

### Measurement of regional infection level of COVID-19

The infection level in the region where participants lived was measured by the incidence rate for the entire period since the pandemic was declared (per 1000 population), the number of people infected for the entire period, the incidence rate for one month before the survey (per 1000 population), and the number of people infected over the one month.

### Assessment of other covariates

Covariates included demographic and socioeconomic factors including age, sex, marital status, household income, educational background, job type, smoking status, and the number of employees in the workplace. Age was used as a continuous variable. Marital status was categorized in three groups: currently married, divorced or widowed, and never married. Annual equivalent household income was categorized into three groups: 470000-2650000 Japanese yen (JPY), 2650000-4500000 JPY, and 4590000-10500000 JPY. Educational background was categorized in three groups: graduated from junior high school; high school; and university, graduate school, vocational school, or junior college. Job type was categorized in three groups: mainly desk work (clerical or computer work), mainly work involving interpersonal communication (customer service, sales, selling, etc.), and mainly labor (physical work, nursing care, etc.). The number of employees in the workplace was classified into four categories: 1-29, 30-99, 100-999, and ≤1000.

### Statistical analysis

We show the participants’ demographic information using counts and percentages (Table 1). We compared seven representative NPIs by the four regions using one-way analysis of variance (Table 2).

**Table 1.** Characteristics of the subjects by area according to incidence rate of COVID-19

|  | Area according to incidence rate of COVID-19<sup>a</sup> |  |  |  |
| --- | --- | --- | --- | --- |
|  | region 1<br>(n=5342) | region 2<br>(n=5450) | region 3<br>(n=5334) | region 4<br>(n=10910) |
| Age, mean | 46.5 (10.7) | 45.8 (10.8) | 47.1 (10.5) | 47.8 (10.3) |
| Sex, male (%) | 2709 (50.7%) | 2766 (50.8%) | 2725 (51.1%) | 5614 (51.5%) |
| Marriage status |  |  |  |  |
| Currently married | 3022 (56.6%) | 3211 (58.9%) | 2999 (56.2%) | 5797 (53.1%) |
| Divorced or widowed | 586 (11.0%) | 588 (10.8%) | 575 (10.8%) | 1094 (10.0%) |
| Never married | 1734 (32.5%) | 1651 (30.3%) | 1760 (33.0%) | 4019 (36.8%) |
| Annual equivalent household income (JPY) |  |  |  |  |
| 470000-2650000 | 1996 (37.4%) | 1764 (32.4%) | 1893 (35.5%) | 3341 (30.6%) |
| 2650000-4500000 | 1786 (33.4%) | 1927 (35.4%) | 1774 (33.3%) | 3195 (29.3%) |
| 4590000-10500000 | 1560 (29.2%) | 1759 (32.3%) | 1667 (31.3%) | 4374 (40.1%) |
| Educational background |  |  |  |  |
| Junior high | 62 (1.2%) | 83 (1.5%) | 77 (1.4%) | 146 (1.3%) |
| High school | 1796 (33.6%) | 1450 (26.6%) | 1483 (27.8%) | 2224 (20.4%) |
| University, graduate school, vocational school,<br>junior college | 3484 (65.2%) | 3917 (71.9%) | 3774 (70.8%) | 8540 (78.3%) |
| Job type |  |  |  |  |
| Mainly desk work (clerical or computer work) | 2689 (50.3%) | 2684 (49.2%) | 2626 (49.2%) | 5469 (50.1%) |
| Jobs mainly involving interpersonal communication<br>(customer service, sales, selling, etc.) | 1287 (24.1%) | 1315 (24.1%) | 1304 (24.4%) | 3021 (27.7%) |
| Mainly labor (physical work, nursing care, etc.) | 1366 (25.6%) | 1451 (26.6%) | 1404 (26.3%) | 2420 (22.2%) |
| Current smoker, % | 1410 (26.4%) | 1302 (23.9%) | 1386 (26.0%) | 2906 (26.6%) |
| Number of employees in the workplace |  |  |  |  |
| 1-29 | 1209 (22.6%) | 1138 (20.9%) | 1257 (23.6%) | 2561 (23.5%) |
| 30-99 | 1613 (30.2%) | 1444 (26.5%) | 1377 (25.8%) | 2506 (23.0%) |
| 100-999 | 1403 (26.3%) | 1574 (28.9%) | 1424 (26.7%) | 2752 (25.2%) |
| ≤1000 | 1117 (20.9%) | 1294 (23.7%) | 1276 (23.9%) | 3091 (28.3%) |
| Incidence of COVID-19 for the whole period (per 1000),<br>median (IQR) | 0.28<br>(0.20-0.34) | 0.51<br>(0.51-0.55) | 0.79<br>(0.66-0.88) | 1.91<br>(1.54-3.12) |
| Number of people infected with COVID-19<br>for the whole period, median (IQR) | 396<br>(211, 448) | 1053<br>(507, 1073) | 1822<br>(974, 2413) | 14427<br>(9309, 27500) |
| Incidence of COVID-19 in the month<br>before the survey (per1000), median (IQR) | 0.89<br>(0.67-0.14) | 0.26<br>(0.15-0.32) | 0.32<br>(0.25-0.44) | 0.74<br>(0.59-1.06) |
| Number of people infected with COVID-19<br>in the month before the survey, median (IQR) | 148<br>(79, 184) | 447<br>(124, 501) | 865<br>(403, 1193) | 5596<br>(3664, 9851) |

<sup>a</sup> Region 1: Fukushima, Yamaguchi, Aomori, Ehime, Yamagata, Nagasaki, Iwate, Tokushima, Shimane, Kagawa, Niigata, Tottori, and Akita prefectures.
Region 2: Nagano, Saga, Tochigi, Oita, Toyama, Okayama, and Fukui prefectures.
Region 3: Gunma, Ishikawa, Gifu, Kumamoto, Ibaraki, Miyagi, Hiroshima, Shiga, Mie, Kochi, Shizuoka, Wakayama, Miyazaki, Yamanashi, and Kagoshima prefectures.
Region 4: Tokyo, Kanagawa, Saitama, Chiba, Okinawa, Osaka, Hokkaido, Aichi, Hyogo, Fukuoka, Kyoto, and Nara prefectures.

**Table 2.** Non-pharmaceutical interventions by area according to incidence rate of COVID-19

|  | total<br>(n=27036) | Area according to incidence rate of COVID-19<sup>a</sup> |  |  |  | p for trend <sup>b</sup> |
| --- | --- | --- | --- | --- | --- | --- |
|  |  | region 1<br>(n=5342) | region 2<br>(n=5450) | region 3<br>(n=5334) | region 4<br>(n=10910) |  |
| Wearing a mask in public | 23308 (86.2%) | 4483 (83.9%) | 4736 (86.9%) | 4624 (86.7%) | 9465 (86.8%) | <0.001 |
| Washing hands after using the bathroom | 23065 (85.3%) | 4513 (84.5%) | 4661 (85.5%) | 4569 (85.7%) | 9322 (85.4%) | 0.176 |
| Disinfecting hands with alcohol sanitizers when going indoors | 15014 (55.5%) | 2850 (53.4%) | 2968 (54.5%) | 2950 (55.3%) | 6246 (57.3%) | <0.001 |
| Gargling when returning home | 13767 (50.9%) | 2393 (44.8%) | 2462 (45.2%) | 2718 (51.0%) | 6194 (56.8%) | <0.001 |
| Opening windows and doors for ventilation | 12155 (45.0%) | 2107 (39.4%) | 2259 (41.4%) | 2462 (46.2%) | 5327 (48.8%) | <0.001 |
| Disinfecting or washing hands after touching frequently-touched surfaces such as doorknobs or railings | 10048 (37.2%) | 1759 (32.9%) | 1975 (36.2%) | 1991 (37.3%) | 4323 (39.6%) | <0.001 |
| Carrying alcohol sanitizers when going out | 8389 (31.0%) | 1464 (27.4%) | 1655 (30.4%) | 1664 (31.2%) | 3606 (33.1%) | <0.001 |

<sup>a</sup> Area was divided according to incidence rate of COVID-19 for the whole period (per 1000).
Median of incidence rates were 0.28 for region 1, 0.51 for region 2, 0.79 for region 3, and 1.91 for region 4.
<sup>b</sup> P-values were derived from the nonparametric test for trend.

Age-sex and multivariate adjusted odds ratios (ORs) of incidence rate of COVID-19 in areas associated with each NPI, defined by those who answered “always do”, were estimated with a multilevel logistic model and nested by area of residence (cities, towns and villages). In the multilevel model, the incidence rate of COVID-19 in an area was used as an area-level factor. The multivariate model was adjusted for age, sex, marital status, education, job type, annual equivalent household income, smoking status, and number of employees in the workplace. A p value less than 0.05 was considered statistically significant. All analyses were conducted using Stata (Stata Statistical Software: Release 16; StataCorp LLC, TX, USA).

## Results

Table 1 shows basic characteristics of the participants. Regions are classified by the incidence of COVID-19 for the entire period since the pandemic was declared (per 1000). Region 4 had the highest infection rate, at 1.91, followed by region 3 (0.79), region 2 (0.51), and region 1 (0.28). Region 4 includes the Tokyo metropolitan area, which has more urban lifestyle characteristics than the other three regions, such as more single people, higher household incomes, higher education levels, more hospitality workers, fewer manual laborers, and more workers in large companies.

Table 2 shows the implementation status of the seven NPIs by region according to the incidence rate of COVID-19. Among the interventions, wearing masks in public places had the highest overall implementation rate, at 86.2%, followed in decreasing order by washing hands after using the bathroom (85.3%), disinfecting hands with alcohol sanitizers when going indoors (55.5%), gargling when returning home (50.9%), opening windows and doors for ventilation (45.0%), disinfecting or washing hands after touching frequently touched surfaces (37.2%), and carrying alcohol sanitizers when going out (31.0%). Six of the NPIs showed a trend in implementation according to area incidence rate of COVID-19 (all ps <0.001); the exception was handwashing after using the bathroom

Table 3 shows the multivariate analyses of the implementation of the seven NPI items. Compared to the region with the lowest infection rate (region 1), the odds ratio for the region with the highest rate (region 4) was 1.24 (95%CI 1.10-1.40, p<0.001) for wearing a mask in public, 1.08 (95%CI 0.97-1.20, p=0.157) for washing hands after using the bathroom, 1.17 (95%CI 1.01-1.35, p=0.031) for disinfecting hands with alcohol sanitizers when going indoors, 1.54 (95%CI 1.31-1.82, p<0.001) for gargling when returning home, 1.45 (95%CI 1.20-1.75, p<0.001) for opening windows and doors for ventilation, 1.33 (95%CI 1.18-1.51, p<0.001) for disinfecting or washing hands after touching frequently touched surfaces, and 1.32 (95%CI 1.17-1.49, p<0.001) for carrying alcohol sanitizers when going out. Five items showed statistically significant trends with regional infection levels: disinfecting hands with alcohol sanitizers when going indoors (p for trend=0.030), gargling when returning home (p <0.001), opening windows and doors for ventilation (p <0.001), disinfecting or washing hands after touching frequently touched surfaces (p <0.001), and carrying alcohol sanitizers when going out (p <0.001). The two NPI items showing no significant trend with regional infection level were wearing a mask in public and washing hands after using the bathroom. Multivariate adjustment did not change any trends.

**Table 3.** Odds ratios of area incidence rate of COVID-19 associated with non-pharmaceutical interventions

|  | Area according to incidence rate of COVID-19<sup>a</sup> |  |  |  |  |  |  |  |  |
| --- | --- | --- | --- | --- | --- | --- | --- | --- | --- |
|  | region 2<br>(n=5450) |  |  | region 3<br>(n=5334) |  |  | region 4<br>(n=10910) |  |  |
|  | OR<sup>b</sup> | 95% CI | p | OR<sup>b</sup> | 95% CI | p | OR<sup>b</sup> | 95% CI | p |
| Wearing a mask in public |  |  |  |  |  |  |  |  |  |
| model 1<sup>c</sup> | 1.26 | 1.10-1.45 | 0.001 | 1.25 | 1.09-1.42 | 0.001 | 1.24 | 1.10-1.40 | <0.001 |
| model 2<sup>d</sup> | 1.23 | 1.07-1.41 | 0.004 | 1.23 | 1.08-1.40 | 0.002 | 1.17 | 1.03-1.32 | 0.013 |
| Washing hands after the bathroom |  |  |  |  |  |  |  |  |  |
| model 1<sup>c</sup> | 1.09 | 0.97-1.24 | 0.143 | 1.10 | 0.98-1.24 | 0.103 | 1.08 | 0.97-1.20 | 0.157 |
| model 2<sup>d</sup> | 1.08 | 0.96-1.22 | 0.236 | 1.09 | 0.97-1.23 | 0.158 | 1.01 | 0.91-1.13 | 0.775 |
| Disinfecting hands with alcohol sanitizers when going indoors |  |  |  |  |  |  |  |  |  |
| model 1<sup>c</sup> | 1.03 | 0.87-1.22 | 0.715 | 1.11 | 0.96-1.28 | 0.147 | 1.17 | 1.01-1.35 | 0.031 |
| model 2<sup>d</sup> | 1.01 | 0.85-1.19 | 0.940 | 1.10 | 0.95-1.28 | 0.194 | 1.14 | 0.98-1.33 | 0.081 |
| Gargling when returning home |  |  |  |  |  |  |  |  |  |
| model 1<sup>c</sup> | 0.96 | 0.80-1.17 | 0.714 | 1.29 | 1.10-1.52 | 0.002 | 1.54 | 1.31-1.82 | <0.001 |
| model 2<sup>d</sup> | 0.94 | 0.78-1.15 | 0.564 | 1.28 | 1.09-1.51 | 0.003 | 1.49 | 1.26-1.76 | <0.001 |
| Opening windows and doors for ventilation |  |  |  |  |  |  |  |  |  |
| model 1<sup>c</sup> | 1.05 | 0.85-1.31 | 0.633 | 1.32 | 1.1-1.59 | 0.003 | 1.45 | 1.2-1.75 | <0.001 |
| model 2<sup>d</sup> | 1.04 | 0.83-1.29 | 0.741 | 1.32 | 1.1-1.58 | 0.003 | 1.41 | 1.17-1.70 | <0.001 |
| Disinfecting or washing hands after touching frequently-touched surfaces such as doorknobs or railings |  |  |  |  |  |  |  |  |  |
| model 1<sup>c</sup> | 1.14 | 0.99-1.31 | 0.069 | 1.23 | 1.09-1.40 | 0.001 | 1.33 | 1.18-1.51 | <0.001 |
| model 2<sup>d</sup> | 1.12 | 0.97-1.29 | 0.123 | 1.22 | 1.08-1.39 | 0.002 | 1.30 | 1.15-1.48 | <0.001 |
| Carrying alcohol sanitizers when going out |  |  |  |  |  |  |  |  |  |
| model 1<sup>c</sup> | 1.12 | 0.98-1.29 | 0.106 | 1.21 | 1.06-1.37 | 0.003 | 1.32 | 1.17-1.49 | <0.001 |
| model 2<sup>d</sup> | 1.11 | 0.96-1.27 | 0.163 | 1.20 | 1.06-1.37 | 0.004 | 1.30 | 1.15-1.48 | <0.001 |

<sup>a</sup> Area was divided according to the incidence rate of COVID-19 for the whole period (per 1000). Median incidence rates were 0.28 for region 1, 0.51 for region 2, 0.79 for region 3, and 1.91 for region 4.
<sup>b</sup> The reference group is region 1 (n=5342).
<sup>c</sup> Model 1 adjusted for age and sex.
<sup>d</sup> Model 2 adjusted for age, sex, marital status, education, job type, annual equivalent household income, smoking status, and number of employees in the workplace.

## Discussion

This study revealed that people living in areas with higher levels of infection were more likely to engage in various non-pharmaceutical interventions. It has been reported that individual characteristics such as sociodemographic factors (6–8), sources of information (9) and psychological factors (7,8,10,11) are associated with non-pharmaceutical behaviors. However, in this study, the association between community infection level and preventive behaviors remained robust even after adjusting for individual factors. The results suggest that the implementation of NPIs is influenced by the regional context.

There are at least three possible reasons why the regional infection level may affect the implementation of NPIs. First, the higher the level of infection in a region, the more people’s fear of infection and their risk perception is likely to be affected, which in turn will lead to greater engagement with NPIs. Japanese people trust local government information more than other sources of information (9), and are relatively knowledgeable about the status of infection in their communities. Second, people living in areas with higher levels of infection will encounter more social situations that require NPIs, such as workplaces, restaurants, and public facilities. Third, majority synching bias (15) may impact people’s implementation of NPIs; this bias is the idea that it is safe to act in the way that people around you are acting and has been demonstrated in a study of factors associated with decision-making regarding evacuation during a disaster (15). Moreover, a recent report found that the predominant reason for wearing a mask during a pandemic among Japanese people was that wearing a mask was a social norm, whereas the original purpose - preventing personal infection and transmitting it to others - was less important (16). Majority synching bias is likely to be enhanced as the number of people performing NPIs increases in areas with high infection levels.

Among the NPIs studied here, some showed a dose-response relationship with the local infection level, whereas others – specifically, wearing masks and hand-washing after using the bathroom – did not. We assume that the latter two were not associated with the local infection level because their implementation levels were high, at about 85%, regardless of infection level. In Japan, wearing masks has been widely practiced during epidemics of common colds, influenza, seasonal allergies, and infectious gastroenteritis, even since before the current global pandemic (17). A 2015 survey by the Consumer Affairs Agency, a Japanese government agency, reported that about 85% of respondents said they washed their hands after using the bathroom (18). It is likely that the lack of a dose-response relationship in relation to COVID-19 and community infection levels is due to Japanese people being accustomed to wearing masks and washing their hands after using the bathroom, which start with hygiene education in childhood.

In contrast, increasing ventilation, carrying alcohol, and hand washing after contacting surfaces were found to have a dose-response relationship with community infection levels. These were newly recommended approaches for helping to prevent the spread of COVID-19. In previous infectious disease epidemics, carrying alcohol-based disinfectant and washing or disinfecting hands after touching frequently handled doorknobs and handrails were not common prevention behaviors. In view of the widespread recognition that alcohol disinfection is effective against SARS-Cov2 (19,20) and that COVID-19 is contact-transmissible (21), it is likely that such behaviors were reinforced as the infection spread, especially in regions with higher infection rates.

Although ventilating rooms is also widely acknowledged as a measure for preventing infection, it has not been routinely adopted because it causes the room temperature to drop during the winter, and many work environments cannot be ventilated appropriately due to the structure of buildings. During this pandemic, avoiding the “Three Cs”-closed spaces with poor ventilation, crowded places, and close-contact settings, has been strongly recommended, especially in Japan (5). As many people are aware that ventilation is highly effective in preventing COVID-19 infection, it is not surprising that this NPI is more commonly performed in regions with higher infection rates.

In Japan, people are taught from a young age that gargling is a preventive measure against infectious diseases, starting in schools and households. Gargling is practiced by many people during infectious disease epidemics. However, in the present survey it emerged that gargling was not so widespread, with a practice prevalence of about 50%. Although gargling may be effective in preventing upper respiratory tract infections in healthy adults (22), it is not recommended in the CDC guidelines for infection prevention (4) or even in the Japanese guidelines for preventive measures against COVID-19 (5). Because of their belief in gargling’s effectiveness, people in areas with higher levels of community infection may be more concerned about possible infection while away from home, and therefore practice more proactive gargling behavior.

The present results suggest that individual adherence to NPIs depends not only on individual characteristics, but also on the contextual effect of local infection level. Considering that the waxing and waning of infection will be repeated in the future, it is important for people to actively implement NPIs even in areas with low infection levels, where organized campaigns to share information about risks and encourage preventive actions may be effective. In addition, although performing NPIs is an individual initiative, the workplace provides an opportunity to use chains of command and require individuals to take active measures to prevent infection.

There are several limitations in this study. First, we conducted a survey of full-time workers only; we did not include part-time workers, housewives, the elderly, or people under 15 years of age. Accordingly, the results do not present a complete picture of NPI practices in the community; some reports show different rates of implementation of preventive measures in different age groups (6). Second, given the cross-sectional nature of the study, it is not possible to assign causation. In other words, it is not possible to compare rates of NPI implementation before and after the increase in COVID-19 infection rate in a given region. Therefore, we cannot conclude that the rate of implementation has increased because of the increase in infectious rate. Third, we used a simple question to measure NPI adherence; details about frequency and timing remain unknown.

To prevent the spread of COVID-19, performing NPIs is essential, along with organizational efforts such as emergency declarations and lockdowns. In this study of Japanese workers, NPIs were more prevalent in regions with higher incidence rates, suggesting that implementation was affected not only by personal attributes but also by the contextual effects of local infection level. In particular, for areas where the implementation is not widespread and the level of infection is low, we propose that environmental interventions such as campaigns to promote awareness of NPIs may be beneficial.

- What is already known on this subject?
  ➢ Non-pharmaceutical interventions have been recommended as a measure to prevent or slow the spread of COVID-19.
  ➢ Adherence to individual NPIs is influenced by sociodemographic factors, psychological factors, and information sources.
- What this study adds?
  ➢ In Japan, people living in areas with higher levels of infection were more likely to engage in non-pharmaceutical interventions.
  ➢ Implementation was affected not only by personal attributes but also by the contextual effects of local infection level.
  ➢ Environmental interventions such as campaigns to promote awareness of NPIs may be beneficial in areas where implementation is not widespread and the level of infection is low.

## Data Availability

Not applicable

## Acknowledgements

This study was supported and partly funded by the University of Occupational and Environmental Health, Japan; General Incorporated Foundation (Anshin Zaidan); The Development of Educational Materials on Mental Health Measures for Managers at Small-sized Enterprises; Health, Labour and Welfare Sciences Research Grants; Comprehensive Research for Women’s Healthcare (H30-josei-ippan-002); Research for the Establishment of an Occupational Health System in Times of Disaster (H30-roudou-ippan-007), Research for AIDS Policy (JPMP 20 HB 1004), and scholarship donations from Chugai Pharmaceutical Co., Ltd., the Collabo-Health Study Group, and Hitachi Systems, Ltd.

The current members of the CORoNaWork Project, in alphabetical order, are Yoshihisa Fujino (present chairperson of the study group), Akira Ogami, Arisa Harada, Ayako Hino, Hajime Ando, Hisashi Eguchi, Kazunori Ikegami, Kei Tokutsu, Keiji Muramatsu, Koji

Mori, Kosuke Mafune, Kyoko Kitagawa, Masako Nagata, Mayumi Tsuji, Ning Liu, Rie Tanaka, Ryutaro Matsugaki, Seiichiro Tateishi, Shinya Matsuda, Tomohiro Ishimaru, and Tomohisa Nagata. All members are affiliated with the University of Occupational and Environmental Health, Japan.

## Disclosure

### Ethical approval

This study was approved by the ethics committee of the University of Occupational and Environmental Health, Japan(reference No. R2-079 and R3-006). **Informed Consent:** Informed consent was obtained in the form of the website.

### Conflict of Interest

The authors declare no conflicts of interest associated with this manuscript.

## Notes

### Competing Interest Statement

The authors have declared no competing interest.

### Author Declarations

This study was approved by the ethics committee of the University of Occupational and Environmental Health, Japan (reference No. R2-079 and R3-006）.

